# Risk Stratification of Non-Missense *KCNH2* Variants Using Deep Generative Models Pretrained on Public Data

**DOI:** 10.64898/2026.11.26.26368681

**Authors:** Takeshi Mitani, Hirohiko Kohjitani, Yuji Okamoto, Takanori Aizawa, Koichi Kato, Takeru Makiyama, Seiko Ohno, Minoru Horie, Yasushi Okuno, Koh Ono

**Affiliations:** Department of Gastroenterology and Hepatology, Kurashiki Central Hospital, Kurashiki, Japan; Department of Artificial Intelligence in Healthcare and Medicine, Graduate School of Medicine Kyoto University, Kyoto, Japan; Department of Cardiovascular Medicine, Kyoto University, Kyoto, Japan; Center for Biosystems Dynamics Research, RIKEN, Kobe, Japan; Department of Cardiovascular Medicine, Shiga University of Medical Science, Otsu, Japan; Medical Genome Center, National Cerebral and Cardiovascular Center, Suita, Japan

**Keywords:** Long QT syndrome, *KCNH2*, Non-missense variant, Risk stratification, Variational autoencoder, Survival analysis

## Abstract

Long QT syndrome type 2 (LQT2) caused by *KCNH2* variants is associated with life-threatening arrhythmias, but prognostic stratification of non-missense variants remains limited. We developed a deep generative model pretrained on 1,955 publicly available *KCNH2* variants and fine-tuned on 403 LQT2 patients. Clustering of non-missense variants in the learned latent space yielded two prognostically distinct clusters (P=0.003). Variants assigned to the high-risk cluster were dominant in specific domains. This approach identifies subgroups with distinct observed outcomes among carriers of non-missense *KCNH2* variants, providing a basis for more individualized risk stratification.

## Introduction

Congenital long QT syndrome (LQTS) is an inherited cardiac channelopathy characterized by delayed ventricular repolarization, predisposing patients to life-threatening ventricular arrhythmias and sudden cardiac death at a young age; accurate risk stratification remains a critical unmet need in clinical practice. *KCNH2*, the causative gene of LQT2, encodes the hERG/Kv11.1 channel responsible for the rapid delayed rectifier potassium current, and loss of its function delays repolarization and prolongs the QT interval(1).

Among *KCNH2* variant carriers, prognostic stratification by variant type and location has advanced primarily for missense variants, in which those in the transmembrane pore region confer a high risk of arrhythmic events(2,3). In contrast, non-missense variants have been treated as a single prognostic group: they have been reported to show no clear risk difference from missense variants or even lower risk, whereas in the C-terminal region (after residue 657) they carry a higher risk than missense variants(2,4). These conflicting findings argue against a single non-missense category, yet stratification within non-missense variants remains unestablished because of their biological heterogeneity and the scarcity of variants linked to prognostic clinical data.

To address this, we applied a joint deep generative framework combining a VAE(5) and DeepSurv(6). Adapting a scheme from prior work in which the two were trained separately(7), we applied a 2 step learning which leveraged unsupervised pretraining on large-scale unlabeled sequence data to compensate for limited clinical labels and extract variant-type-independent representations to investigate prognostic stratification in patients with LQT2 harboring non-missense *KCNH2* variants.

## Methods

### 1. Study population

In this retrospective, multicenter cohort study, we used existing institutional datasets of patients referred to Shiga University of Medical Science or Kyoto University Graduate School of Medicine for genetic evaluation between 1996 and 2021.

Eligible patients carried rare *KCNH2* variants but no rare variants in other known LQTS-related genes and had available sex and outcome data. Genetic testing and variant definition were performed as previously described by Aizawa et al(4). This clinically annotated dataset was used for fine-tuning. For pretraining, a variant-only dataset was extracted from dbSNP, National Center for Biotechnology Information (NCBI); all variant types except synonymous variants were included.

### 2. Clinical outcomes

The primary endpoint was time from birth to the first arrhythmic event (AE), defined as syncope, life-threatening ventricular arrhythmia, or death. Participants were censored at the earliest of last clinical contact, initiation of pharmacological therapy, or age 40 years.

### 3. Model

#### 3-1. Data and Domain Mapping

Variant-specific amino acid sequences based on GRCh38.p14 were aligned to a fixed length with MAFFT with default settings. Residues were encoded using eight prespecified AAindex(8) descriptors and gaps by zero vectors; sex was one-hot encoded in two dimensions. For domain analysis, variants were assigned by the first position differing from hERG1a (UniProt Q12809) to the eag/PAS domain (EAGD), N-linker, voltage-sensing domain (VSD), pore domain (PD), C-linker, cyclic nucleotide-binding homology domain (CNBHD), or C-tail. Boundaries were based on the full-length hERG cryo-EM structure(9). Frameshift, nonsense, and splice-site variants were mapped to the first altered position.

We utilized two types of datasets: one for pretraining and the other for fine-tuning. Let *D_i_^pre^* and *_i_^fine^* denote the observed data for patient *i* in the pretraining and fine-tuning stages, respectively.

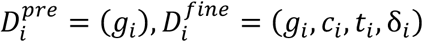

where *g_i_* denotes variant information, *c_i_* denotes sex, *t_i_* denotes follow-up time, and δ*_i_* is the event indicator (1: AE occurred, 0: right-censored).

#### 3-2. Model Architecture

The model comprised a VAE and survival head (Figure 1). The encoder mapped *g_i_* to a latent representation *z_i_*, parameterizing *q***_φ_**(*z_i_*|*g_i_*). The decoder reconstructed *g*_i_ from *z_i_*. The survival head employed a DeepSurv-style Cox proportional hazards model and estimated individual risk η*_i_* from *z_i_* and *c_i_*.

**Figure 1.**
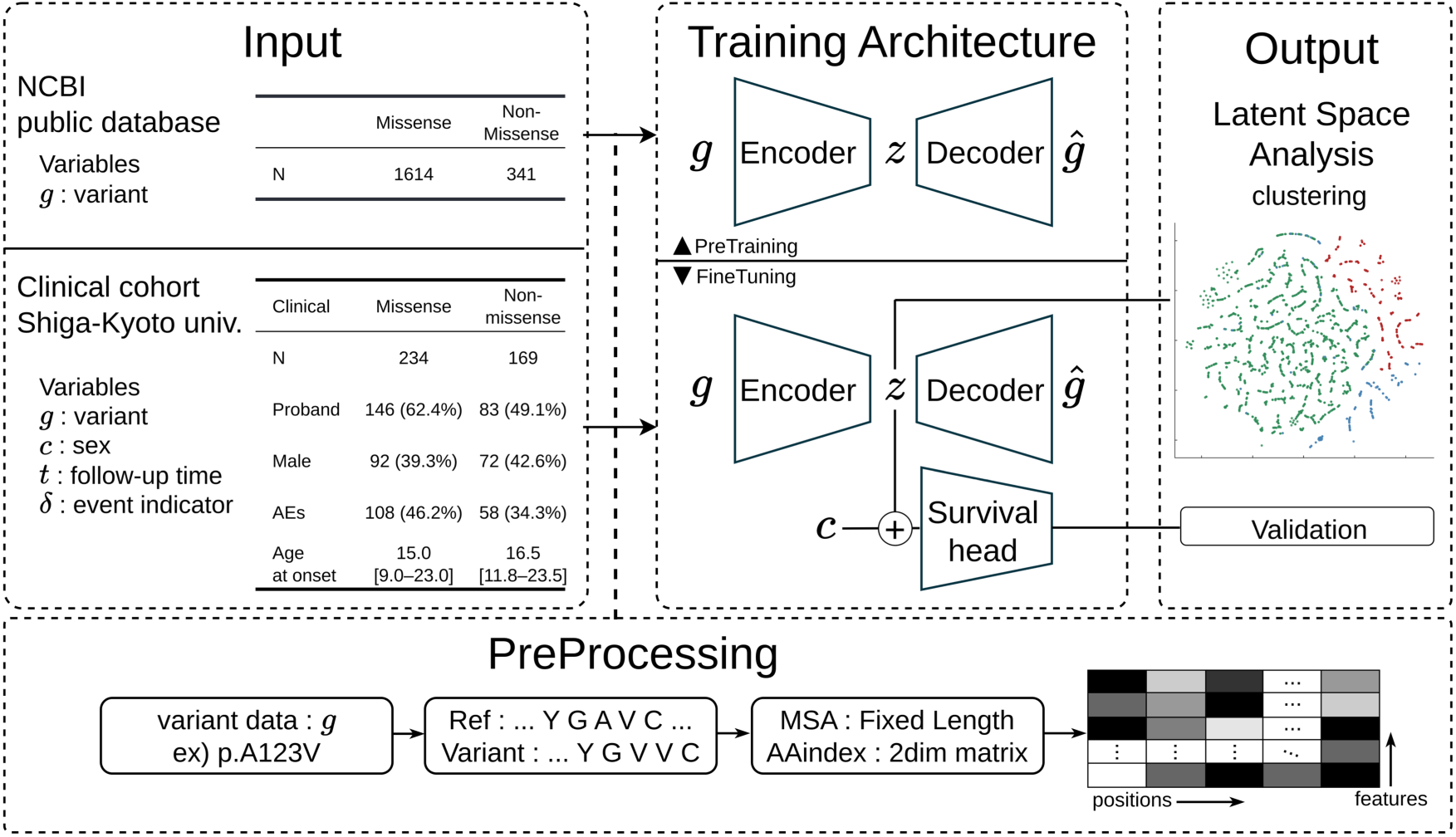
Model architecture. We performed unsupervised pre-training of a variational autoencoder using public *KCNH2* variant records and then fine-tuned the model based on outcomes using a clinical cohort through a survival head. An overview of the public dataset and clinical cohort is provided. Variant sequences were aligned to the reference sequence and encoded by AAindex matrices. The encoder generated latent representations, the decoder reconstructed the input, and the survival head estimated individual risks based on the latent representations and sex.

#### 3-3. Pretraining Objective

In the pretraining stage, the model was optimized without the survival analysis component:

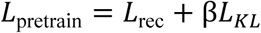

where *L*_pretrain_ denotes the total pretraining loss, *L*_rec_ the reconstruction loss, *L_KL_* the Kullback-Leibler divergence term, and β controls how strongly the latent space is constrained toward a simple normal distribution.

The reconstruction loss was defined using cross-entropy:

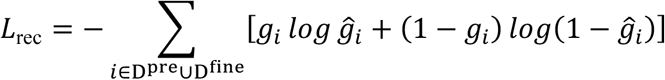

Latent-space regularization used the Kullback–Leibler divergence from the standard normal prior:

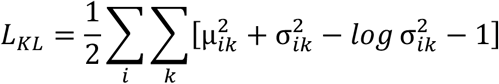

#### 3-4. Fine-Tuning Objective

In fine-tuning, a survival loss term was added to the pretraining objective:

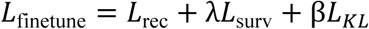

where *L*_finetune_ denotes the fine-tuning loss, *L*_surv_ the survival loss, and λ weights the survival loss.

The survival loss was defined as the negative Cox partial log-likelihood:

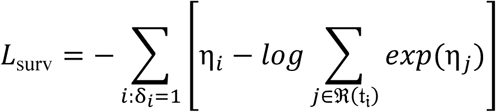

where ℜ(*t_i_*) denotes the risk set at time *t_i_* .

#### 3-5. Latent-Space Analysis

After holdout evaluation, the selected model was refit on the full cohort for exploratory latent-space analysis; *L*_finetune_ was monitored at each epoch with 15-epoch early stopping. Latent representations were clustered using a two-component Gaussian mixture model (GMM). The GMM was fitted to non-missense variant carriers only; missense carriers served as the reference group. Event-free survival was estimated using the Kaplan-Meier method and compared with the log-rank test. Within each domain, cluster proportions (95% Wilson CIs) were compared with those of all other domains using 2×2 chi-square tests. The t-distributed stochastic neighbor embedding (t-SNE) projections were annotated with the Levenshtein distance from the reference sequence.

#### 3-6. Model Setting and Code Availability

Hyperparameters were tuned by grid search with 3-fold cross-validation in the training set. Mean C-index was monitored with early stopping after 15 epochs without improvement. Performance on the prespecified holdout set was assessed using C-index and integrated Brier score (IBS), with standalone DeepSurv as a reference. Proband-only fine-tuning (n=229; 124 AEs) repeated the same procedure. Source code, including the hyperparameter search space, is available at GitHub (https://github.com/mitani111/lqt2-nonmissense-risk).

### 4. Ethics

The study conformed to the Declaration of Helsinki and was approved by the institutional review boards of Shiga University of Medical Science (G2011-128) and Kyoto University Hospital(G1399). Written informed consent was obtained from all participants or their guardians.

## Results

### 1. Study population

The clinical cohort included 403 patients carrying 172 distinct *KCNH2* variants; 166 experienced an AE. Of the 403 patients, 234 (58%) carried missense variants, and AEs were more frequent in missense than in non-missense variant carriers (108 of 234 [46%] vs 58 of 169 [34%]; P=0.023). The public pretraining dataset contained 1,955 *KCNH2* variant records (Figure 1).

### 2. Model performance

In the holdout dataset, the proposed model showed performance similar to standalone DeepSurv: IBS, 0.175 (95% CI, 0.142–0.205) vs 0.175 (0.140–0.206); C-index, 0.566 (0.433–0.678) vs 0.541 (0.436–0.661).

### 3. Latent space analysis

Figure 2A and B show t-SNE projections before and after fine-tuning, respectively, colored by Levenshtein distance from the reference sequence. The GMM assigned 57 of 169 carriers to Cluster A and 112 to Cluster B. Among patients with non-missense variants, event-free survival differed between Cluster A and B (log-rank P=0.003; Figure 2C). Across the 3 groups, Cluster A was comparable to missense variant carriers (P=0.608), whereas Cluster B showed better event-free survival (P=0.002)

**Figure 2.**
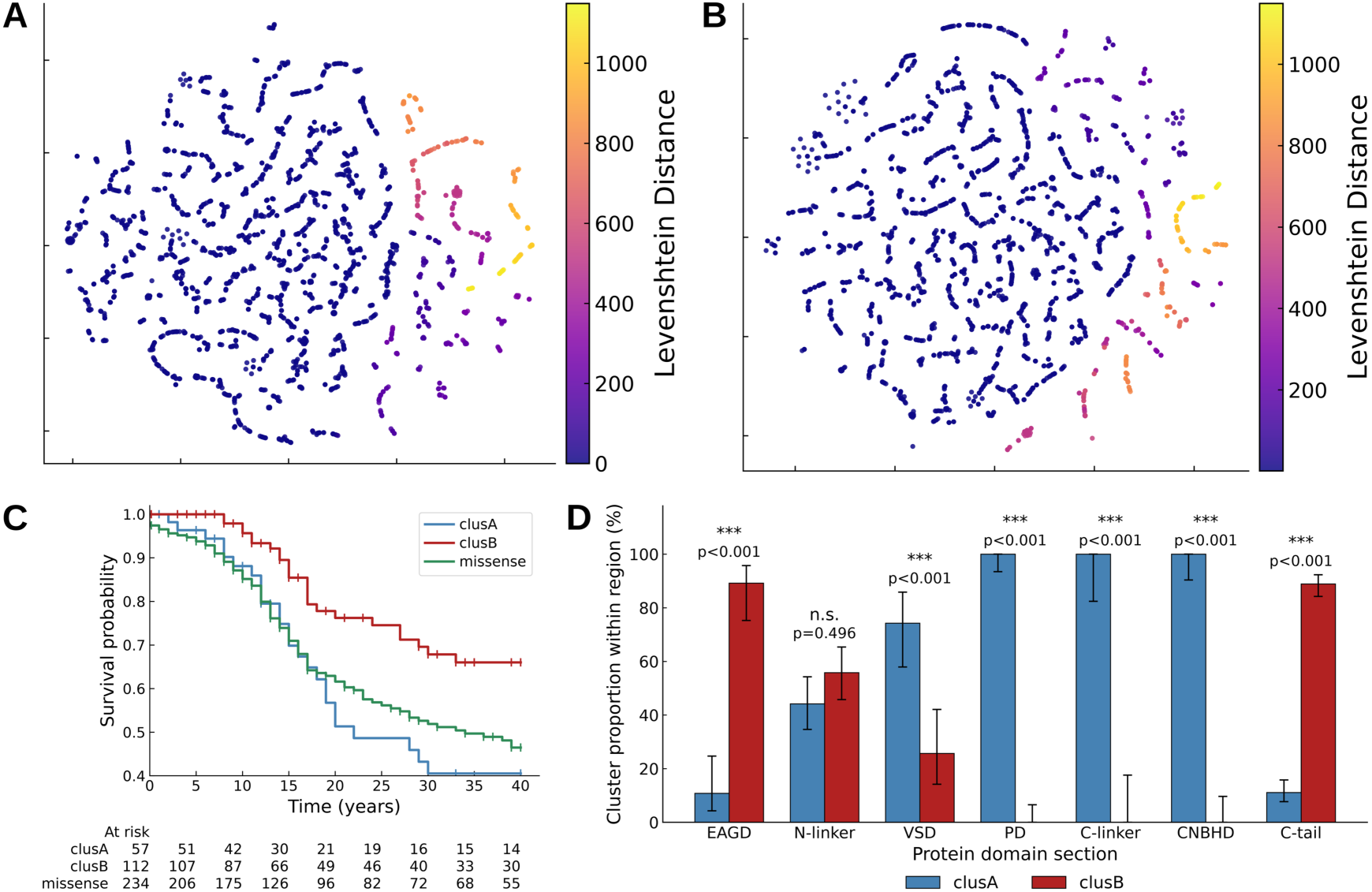
Latent space analysis. (A, B) t-SNE projections before and after fine-tuning, respectively, are colored by Levenshtein distance from the reference KCNH2 sequence. (C) Kaplan-Meier curves show event-free survival among patients with non-missense variants in Clusters A and B and patients with missense variants. (D) Bars show the proportions of non-missense variants assigned to Clusters A and B within each structural domain; error bars indicate 95% Wilson CIs. Chi-square P values compare each domain with all other domains combined (*P<0.05, **P<0.01, ***P<0.001, n.s not significant). Abbreviations: EAGD, eag/PAS domain; N-linker, N-terminal linker; VSD, voltage-sensing domain; PD, pore domain; C-linker, C-terminal linker; CNBHD, cyclic nucleotide-binding homology domain; C-tail, C-terminal tail.

### 4. Domain analysis

Among non-missense variants, cluster membership varied by structural domain (Figure 2D). Cluster A predominated in the VSD, PD, C-linker, and CNBHD, whereas Cluster B predominated in the EAGD and C-tail.

## Discussion

### 1. Effectiveness of Fine-Tuning for Latent Space

We developed a VAE-DeepSurv framework combining unsupervised pretraining on public *KCNH2* variant records with outcome-guided fine-tuning in a clinical LQT2 cohort. Prediction accuracy was comparable to DeepSurv, but integrating prognostic and sequence information in the latent space enabled analysis of prognostic heterogeneity within non-missense variants. In proband-only analysis, model performance peaked at epoch 1, suggesting that learning latent representations that jointly capture sequence and prognostic information may be challenging. The primary latent-space analysis therefore used the full cohort, including probands and relatives.

### 2. LQT2 Risk Stratification

Non-missense variants separated into two latent clusters with different observed prognosis (Figure 2C), and their distribution differed across established structural domains (Figure 2D). This supports a potential interaction between variant type and location but does not establish a domain-based risk rule. Independent validation is required.

### 3. Limitations

Discrimination was modest, with 95% CIs overlapping those of DeepSurv. The small cohort relative to *KCNH2* variant diversity, unmeasured environmental effects, and full-cohort outcome-guided latent analysis may limit generalizability. The clusters also lack mechanistic interpretation; their association with structural domains requires biological and external clinical validation.

## Conclusion

Public-data pretraining followed by clinical fine-tuning generated variant-type-independent representations and identified prognostically distinct latent clusters among non-missense *KCNH2* variants. This framework warrants prospective external validation before clinical application.

## COMPETENCY IN MEDICAL KNOWLEDGE

Non-missense KCNH2 variants are prognostically heterogeneous rather than a single risk category. Outcome-guided latent representations separated non-missense variant carriers into two groups with distinct event-free survival, and the high-risk group was dominant in the VSD, PD, C-linker, and CNBHD, indicating that variant type and location should be considered jointly.

## TRANSLATIONAL OUTLOOK

Deep generative modeling pretrained on public variant data supports a novel, data-driven classification of non-missense *KCNH2* variants that could refine individualized risk assessment for patients with LQT2. External validation in larger cohorts and functional characterization of cluster-defining variants are the next steps toward clinical translation.

## Acknowledgments

The authors used Claude (Anthropic) to improve the English language and readability of this manuscript. This research was supported by the Moonshot Research and Development Program under Grant Number JP (JPMJMS2021), AMED under Grant Number (25he2822017j0001) and JSPS KAKENHI Grant Number JP (JP23K15155).

## Conflict of interest

None declared.

## Data availability

The data underlying this article will be shared on reasonable request to the corresponding author.

## Abbreviations and Acronyms

AE: arrhythmic event
C-index: concordance index
CNBHD: cyclic nucleotide-binding homology domain
EAGD: eag/PAS domain
GMM: Gaussian mixture model
IBS: integrated Brier score
LQT2: long QT syndrome type 2
PD: pore domain
VAE: variational autoencoder
VSD: voltage-sensing domain

---

**CENTRAL ILLUSTRATION:**
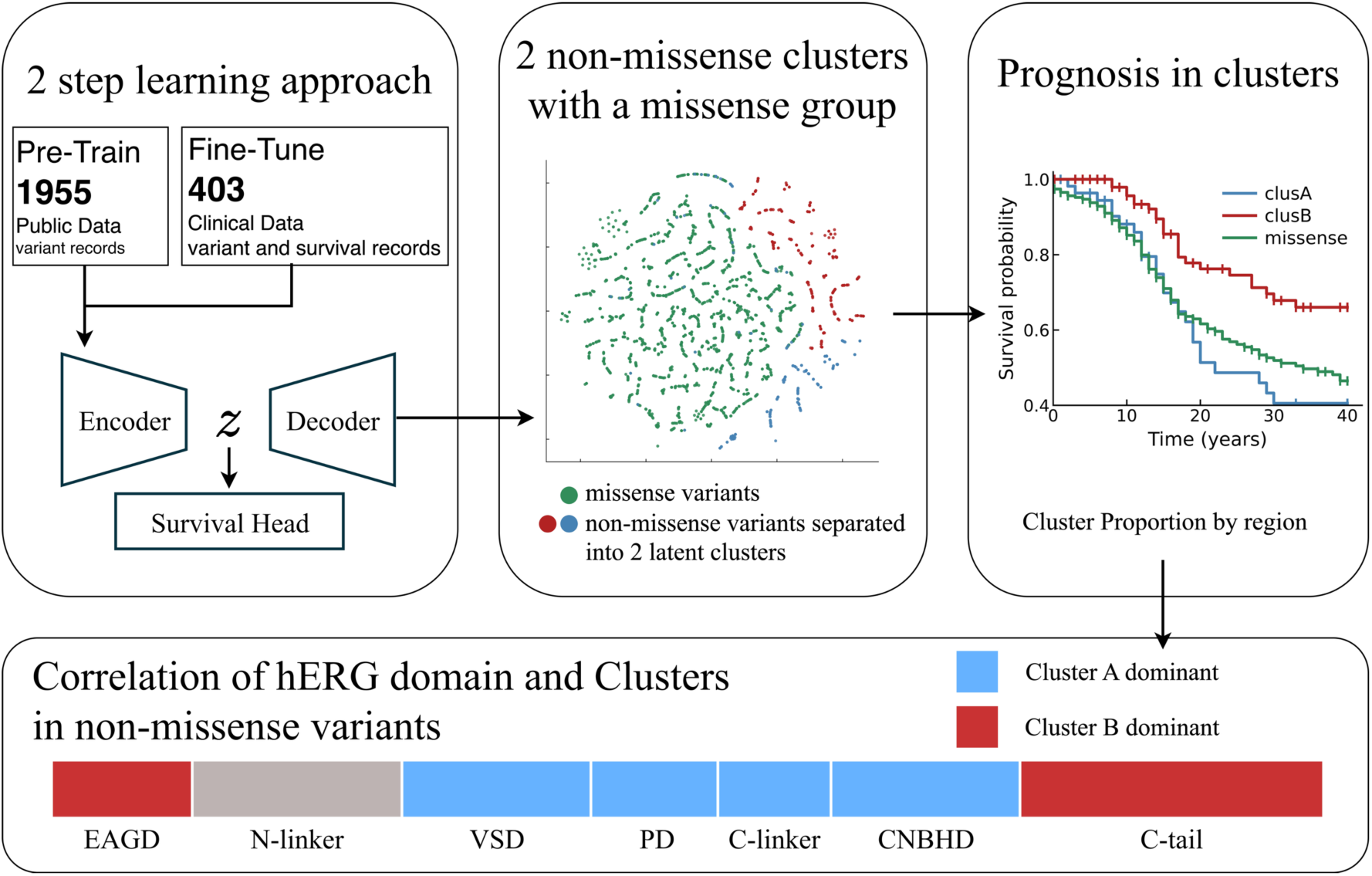
Deep Generative Modeling of Public KCNH2 Data Identifies High-Risk Non-Missense Variants in Long QT Syndrome Type 2. **A** variational autoencoder was pretrained on public *KCNH2* variant cohort and fine-tuned with a survival head on clinical cohort. After fine-tuning, non-missense variants were separated into 2 latent clusters by a Gaussian mixture model. Among patients with non-missense variants, event-free survival differed between Cluster A and B (log-rank P = 0.003). Cluster A was dominant in the VSD, PD, C-linker, and CNBHD, while Cluster B in the EAGD and C-tail. CNBHD = cyclic nucleotide-binding homology domain; PD = pore domain; EAGD = eag/PAS domain; VSD = voltage-sensing domain.

## Notes

### Competing Interest Statement

The authors have declared no competing interest.

### Author Declarations

Ethics IRB of Shiga University of Medical Science and Kyoto University Hospital waived ethical approval for this work.

